# Fertility desires and attitudes toward fertility and childbearing among women of reproductive age

**DOI:** 10.64898/2026.01.07.26343593

**Authors:** Jurczak Anna, Nenko Ilona, Urszula M. Marcinkowska

**Author notes:** Nenko Ilona Correspondence.

## Abstract

**Objectives:** Understanding fertility desires is one of the key components for understanding total fertility rates. The study aims to explore how attitudes towards fertility and childbearing are associated with fertility desires.

**Methods:** A cross-sectional online survey was conducted among 822 childfree, heterosexual women aged between 18 and 35 who were involved in a romantic relationship and were not diagnosed with infertility. The relationship between attitudes to fertility and childbearing and both (1) the desire to have children and (2) the preferred timing of childbearing was analyzed.

**Results:** Women who had a higher score in the Fertility and the child as an important value subscale and Personal awareness and responsibility concerning having a child subscale were more likely to (1) want to have a child and (2) want to have a child sooner (within the next 5 years). Conversely, higher scores on the A child as a barrier subscale were associated with a decreased likelihood of desiring a child and an increased likelihood of planning to have a child later than in the next five years.

**Conclusion:** The findings highlight the role of fertility-related attitudes in shaping reproductive intentions among women of reproductive age.

## Introduction

### Contextual background

Replacement level fertility, which guarantees that a population replaces itself from one generation to the next, is 2.1 children per woman. Yet, the Total Fertility Rate (TFR) in Europe and Poland is considerably lower: 1.53 and 1.16, respectively (GUS, 2024; Statista, 2024). Notably, Poland has been below the replacement level since 1990 (World Data Bank, 2024). As Wilkins (2019) notes, Poland is among the countries with a very low fertility rate, falling far below the threshold for this category, namely 1.6. One of the principal factors contributing to this decline is the increasing average age at which women give birth to their first child in Western, Educated, Industrialised, Rich and Democratic (WEIRD) countries since 1950. In Poland, this average has increased, from 22.7 years in 1990 to 29.0 years in 2023 (GUS, 2024). In addition, the widespread availability of contraception has empowered women to control the timing of the pregnancies, but it is also contributes to postponing childbearing (Wilkins, 2019).

The childbearing postponement decision is influenced by a combination of socio-demographic, contextual and psychological factors (Hashemzadeh et al., 2021; Beaujouan & Toulemon, 2021). Factors such as education and religiosity significantly impact the decision to have children (Hashemzadeh et al., 2021). Religious ties and traditional family models tend to correlate with a higher desire for children, while completing higher education delays this desire (CBOS, 2010; Mills et al., 2011; Bein et al., 2021). A further factor contributing to the low TFR is the perceived economic instability experienced by younger generations (Sobotka, 2017; Suwada, 2019). Young people are confronted with job insecurity and financial instability, which contribute to the perception that parenthood is a challenging prospect. On one hand there is the increasing difficulty of acquiring home ownership, a mortgage (Suwada, 2019), the immediate costs associated with housing and children’s education; and on the other hand, the desire to establish a stable living situation before having children further complicates the readiness to start a family (Bryx et al., 2021). Moreover, the challenges of combining work and parenting responsibilities (e.g. limited access to childcare) are significant factors (Suwada, 2019; Wilkins, 2019). Taken all together, current literature highlights the complex interplay between the socio-demographic and economic factors shaping fertility rates, while also emphasising the role of individual variation in fertility desires.

Fertility desires exert a significant influence on the decision to have a child (Hashemzadeh et al., 2021) and are shaped by a multitude of factors, among them: economic conditions, age, parity, education, health concerns, religiosity, psychological well-being and gender role attitudes (Nitsche & Hayford, 2020; Buber-Ennser & Berghammer, 2021; Hashemzadeh et al., 2021; Golovina et al., 2023). In 2010, among childless Polish citizens aged 18-24, 86% of women and 78% of men wanted children. However, in the 35-39 age group, men were almost twice as likely (47%) as women (28%) to want children (CBOS, 2010). Nevertheless, the latest research showed that in Poland among childfree women aged 18-45, 42% indicated that they do not intend to have children at all. Of the remaining respondents, 34% plan to have children in the next four years or more, while 25% plan to have children within the next three to four years (CBOS, 2023).

A new trend in family planning has recently emerged, namely an increasing number of individuals and couples who consciously do not want to have children at all (Tocchioni et al., 2022). Such a group of people (among those who could become parents) was almost non-existent in previous studies of the Polish population. Abramowska-Kmon et al. (2023) and Mynarska & Brzozowska (2022) found that people who have a negative attitude towards children in general do not actually want to have them. Motivations for remaining childfree differ between genders, with the emotions associated with pregnancy and having an infant being more important for women, and the satisfaction associated with raising and educating a child, and more traditional values being more important for men (Mynarska & Rytel, 2020). However, both genders perceive child-related responsibilities as burdensome and undervalue the emotional aspects of parenthood (Mynarska & Rytel, 2020).

### Study aim

We can observe lowering fertility rates globally, and we know that total fertility is strongly related with fertility desires. Prior research on attitudes towards fertility has concentrated on the positive or negative motivations for parenting, including the enjoyment of pregnancy, childbirth and infancy, traditional parenting, satisfaction with parenting, feelings of being needed and connected, instrumental values, discomfort with pregnancy and childbirth, fears and anxieties about parenting, negative aspects of childcare, and parenting stress (Mynarska & Rytel, 2020). Whereas Söderberg et al. (2013) proposed that an individual’s beliefs and attitudes towards motherhood should also be the focus of investigation, and designed the Attitudes to Fertility and Childbearing Scale (AFCS). The scale captures attitudes towards parenthood on three levels: 1) fertility and the child as an important value, 2) personal awareness and responsibility for having a child, and 3) a child as a barrier. This allows parental attitudes to be examined in relation to three different areas of life, providing a holistic picture of parental motives.

The current study contributes to the theoretical advancement of three extant theories. Firsly, the model under discussion is based on the theoretical framework of the Theory of Planned Behaviour (Ajzen, 1991), which posits that intentions are influenced by attitudes, subjective norms and perceived behavioural control. The AFCS used in this study enabled the exploration of attitudes, and the assessment of personal values, perceived obstacles and a sense of responsibility associated with childbearing. Secondly, Expectancy-Value Theory (Eccles, 1983; Eccles & Wigfield, 2002) posits that an individual’s motivation to engage in a particular behaviour is dependent on the perceived value of the outcome of that behaviour, and on the belief that the individual is capable of achieving that outcome. In the context of this study and the research onto parental intentions, it is important to consider the attitudes and beliefs held by individuals. Thirdly, within the framework of the present study, a final theory to be considered is The Second Demographic Transition (Lesthaeghe, 1995; Van De Kaa, 1987), which posits a decline in fertility in conjunction with shifts in cultural values. These changes are characterised by a shift towards individualism, egalitarianism and self-realisation. Integrating these theories enables understanding of how parental attitudes are embedded in broader socio-cultural contexts that shape reproductive plans.

The present study allows for an exploration of parental attitudes in relation to three distinct domains of life, thereby providing a comprehensive overview of parental motivations. Moreover, socio-demographic factors are also considered. By offering an innovative insights into the relationship between attitudes towards parenting and reproductive intentions this study fills a gap in the literature. Thus, the aim of this study was to investigate how attitudes toward parenthood relate to the desire to have children, both in terms of timing and the overall desire among childfree Polish women. We hypothesised that women who perceived the child as an obstacle were less likely to intend to have offspring in the future and wanted to have them later. In contrast, women who emotionally aligned with the values of childrearing and fertility, and/or exhibited higher personal awareness and responsibility concerning having a child, were more likely to want a child, and they wanted it sooner.

## Methods

### Sample and design

In total 2,639 individuals (18 and 65 years old) participated in a cross-sectional, anonymous online survey between April and July 2021 using the Qualtrics tool. The participants were recruited via social media and online groups. Among other variables (for further details regarding the sample see Golovina et al. (2023) and Marcinkowska et al. (2025), the respondents answered questions related to their socio-demographic status (e.g., age, marital status, socio-economic status), and a Polish adaptation of the Attitudes to Fertility and Childbearing Scale (AFCS; (Kossakowska & Söderberg, 2021). For the purpose of this study, we selected 822 women who met the following criteria: identification as female, heterosexual orientation, age between 18 and 35, involvement in a romantic relationship, lack of children or pregnancy, and lack of any diagnosed infertility (see Table 1 for descriptive statistics of the sample). The study protocol was approved by The Bioethics Committee at Jagiellonian University (opinion number 1072.6120325.2020, obtained on 25/11/2020), and informed consent was obtained from all participants. No financial compensation was provided to participants.

**Table 1.**
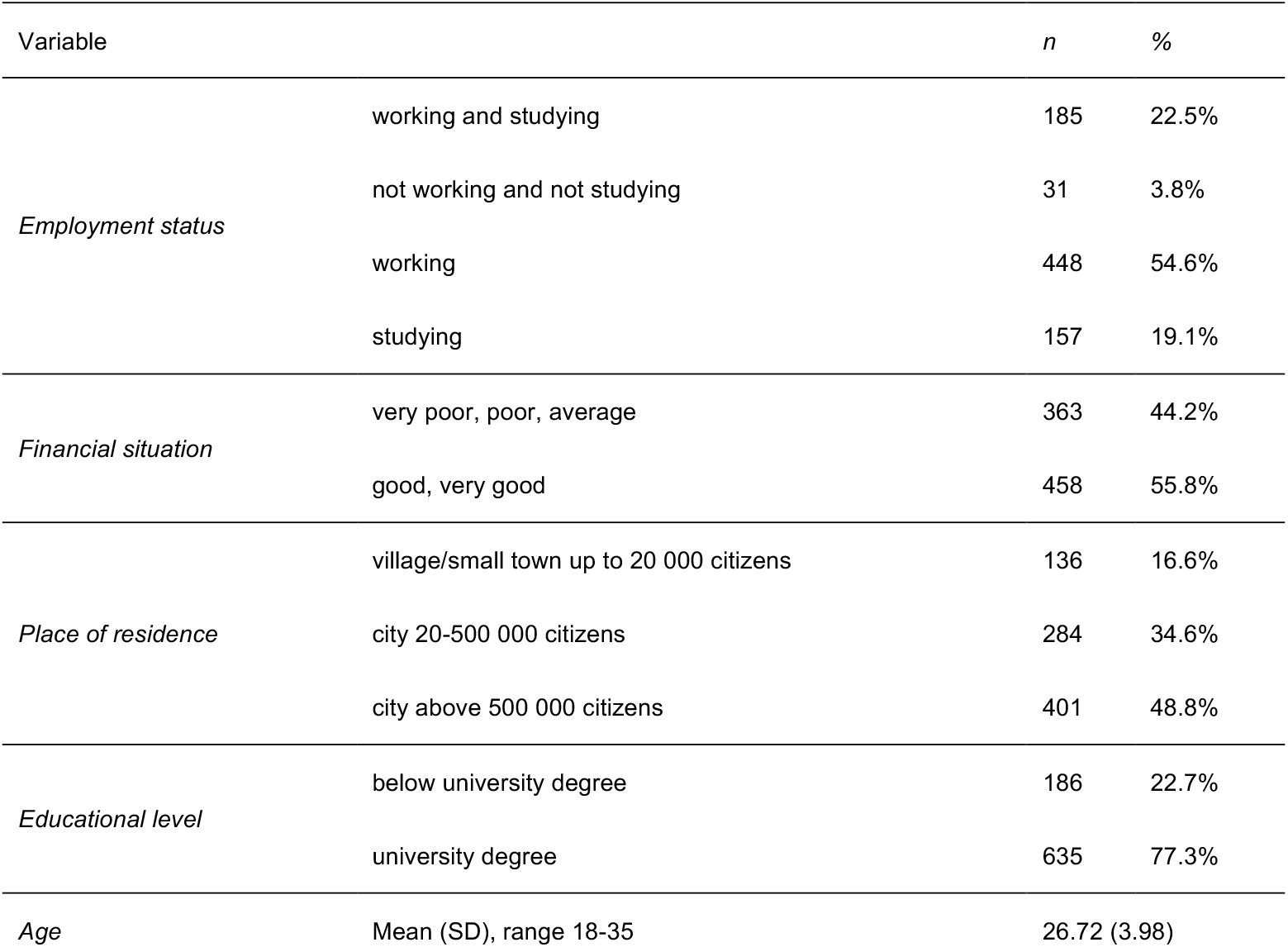
Descriptive statistics.

### Measures

#### Attitudes to Fertility and Childbearing

The Attitudes to Fertility and Childbearing Scale (AFCS) is a 26-item self-report scale using a 5-point Likert scale (1 = “I strongly disagree” to 5 = “I strongly agree”). The scale comprised three subscales: (1) Fertility and the child as an important value (11 items, participant can get 11-55 points), (2) A child as a barrier (10 items, 10-50 points), and (3) Personal awareness and responsibility concerning having a child (5 items, 5-25 points). The higher the score, the more consistent the person’s beliefs were with a given attitude. The AFCS does not have a general score, i.e. subscales had to be analysed separately. The Polish AFCS showed a Cronbach’s alpha reliability coefficient of 0.95 for fertility and the child as an important value, 0.75 for personal awareness and responsibility regarding having a child and 0.85 for a child as a barrier (Kossakowska & Söderberg, 2021).

#### Fertility desires

Firstly, the participants were asked to indicate the number of children they would like to have. Women answered the question: “How many children would you like to have now?”. Secondly, women answered the question: “At what age would you like to have your first child?”. Participants could choose from the following 7 categories: “under 19 years”, “between 20-24 years”, “between 25-29 years”, “between 30-34 years”, “between 35-39 years”, “40 years or older”, and “I do not plan on having children”. Those who selected the last option (i.e. declared that they didn’t plan on having children) were excluded from the analysis of childbearing timing. The difference between the participants’ current age (categorized into 7 categories corresponding to the timing of the childbearing question) and their desired age for having a child was calculated.

#### Socio-demographic characteristics

The questionnaire included questions on the following variables: age, education (levels: primary, vocational, secondary, Bachelor’s, Master’s degree, PhD), employment status (levels: working and studying, not working and not studying, working, studying), perceived financial situation (levels: very poor, poor, average, good, very good), and place of residence (levels: village, small town up to 20 000 citizens, city 20-100 000 citizens, city 100-200 000 citizens, city 200-500 000 citizens, city above 500 000 citizens). Due to the size of the groups, the levels of the variables have been combined (see Table 1 and Statistical Analysis section for details).

### Statistical analysis

The relationship between attitudes towards fertility and the desire to have children, as well as the timing of childbearing was examined using multiple logistic regression models. In the first model about the desire to have children, the raw variable (answers ranging from 0 to 6 children) was dichotomized and was coded as: 0 – women who did not want to have children and 1 – women who indicated a desire to have at least one child. Confounding variables, included: age (continuous), place of residence (0 – village or small town up to 20 000 citizens, 1 – city 20-500 000 citizens, 2 – city above 500 000 citizens), employment status (1 – working and studying; 2 – not working nor studying; 3 – working; 4 – studying), financial situation (0 – very poor, poor or average; 1 – good or very good), and education (0 – below university degree; 1 – university degree). In the second model about the timing of childbearing, the answers were dichotomized and coded as: 0 – women who wanted to have children in the next 5 years and 1 – women who wanted to have children in more than 5 years. Confounding variables were the same as in the first model, with one exception: the age was excluded from this model as a potential confounding variable, because the response variable was based on the participants’ current age. A separate logistic regression model was created for each of the 3 subscales of the AFCS. All analyses were conducted in R v. 2024.04.0+735 (R Core Team, 2024), the *gtsummary* (Sjoberg et al., 2021), the *broom* (Robinson et al., 2024), the *gt* (Iannone et al., 2024) and the *dplyr* (Wickham et al., 2023) packages. The full code is available in the Electronic Supplementary Material (https://doi.org/10.17605/OSF.IO/7KJS5).

## Results

### Desire to have children

#### Fertility and the child as an important value

Women who perceive a child as an important value have increased odds of desire for a child. The odds of desiring a child increase by 20% with each additional point on the “Fertility and the Child as an Important Value” subscale (aOR = 1.20, 95% CI: 1.17 – 1.23), Table 2. This model explains 62.3% of the variance in the desire to have children (Nagelkerke R^2^ = 0.623, *p* < 0.001).

**Table 2.** Fertility desires and attitudes to the childbearing and fertility scale – results from multiple logistic regression models.

| AFCS subscale | Child as an important value<sup>1</sup> |  |  | Personal awareness<sup>2</sup> |  |  | A child as a barrier |  |  |
| --- | --- | --- | --- | --- | --- | --- | --- | --- | --- |
| Characteristic | aOR<sup>3</sup> | 95% CI<sup>4</sup> | p-value<sup>5</sup> | OR<sup>3</sup> | 95% CI<sup>4</sup> | p-value<sup>5</sup> | OR<sup>3</sup> | 95% CI<sup>4</sup> | p-value<sup>5</sup> |
| **AFCS subscale** | 1.20 | 1.17, 1.23 | **<0.001** | 1.78 | 1.65, 1.93 | **<0.001** | 0.83 | 0.80, 0.85 | **<0.001** |
| **Financial situation** |  |  |  |  |  |  |  |  |  |
| *very poor, poor or average* | reference |  |  |  |  |  |  |  |  |
| *good or very good* | 1.33 | 0.88, 2.02 | 0.180 | 1.34 | 0.87, 2.08 | 0.185 | 1.26 | 0.87, 1.82 | 0.219 |
| **Employment status** |  |  |  |  |  |  |  |  |  |
| *neither studying nor working* | reference |  |  |  |  |  |  |  |  |
| *working* | 1.37 | 0.48, 4.12 | 0.564 | 1.08 | 0.33, 3.36 | 0.897 | 1.27 | 0.48, 3.48 | 0.634 |
| *studying and working* | 1.90 | 0.62, 6.13 | 0.269 | 1.20 | 0.35, 3.96 | 0.767 | 1.53 | 0.54, 4.40 | 0.428 |
| *studying or working* | 1.20 | 0.37, 4.00 | 0.760 | 1.00 | 0.28, 3.39 | 0.994 | 1.45 | 0.50, 4.25 | 0.498 |
| **Age (years)** | 0.93 | 0.87, 1.00 | 0.066 | 0.93 | 0.86, 1.00 | 0.057 | 0.86 | 0.80, 0.92 | **<0.001** |
| **Education level** |  |  |  |  |  |  |  |  |  |
| *primary, vocational, secondary* | reference |  |  |  |  |  |  |  |  |
| university degree | 2.18 | 1.21, 3.96 | **0.010** | 2.19 | 1.20, 3.98 | **0.010** | 2.12 | 1.29, 3.50 | **0.003** |
| **Place of residence** |  |  |  |  |  |  |  |  |  |
| *city between 20 000-500 000* | reference |  |  |  |  |  |  |  |  |
| *city above 500 000* | 1.17 | 0.74, 1.85 | 0.497 | 1.09 | 0.68, 1.75 | 0.716 | 1.12 | 0.74, 1.69 | 0.582 |
| *village, town up to 20 000* | 0.89 | 0.46, 1.72 | 0.732 | 1.15 | 0.60, 2.26 | 0.671 | 0.82 | 0.47, 1.41 | 0.465 |
| <sup>1</sup>Fertility and the child as an important value <sup>2</sup>Personal awareness and responsibility concerning having a child, <sup>3</sup>OR = Odds Ratio, <sup>4</sup>CI = Confidence Interval, <sup>5</sup>p-values<0.05 in bold |  |  |  |  |  |  |  |  |  |

#### Personal awareness and responsibility concerning having a child

Women who are more aware and more responsible concerning having a child have increased odds of desire for a child. The odds of desiring a child increase by 78% with each additional point on this subscale (aOR = 1.78, 95% CI: 1.65 – 1.93), Table 2. The model explains 64.1% of the variance in the desire to have children (Nagelkerke R^2^ = 0.641, *p* < 0.001).

#### A child as a barrier

Women who perceive a child as a barrier have decreased odds of desire for a child. The odds of desiring a child decrease by 17% with each additional point on this subscale (aOR = 0.83, 95% CI: 0.80-0.85), Table 2. This model explains 48.6% of the variance (Nagelkerke R^2^ = 0.486, *p* < 0.001).

### Timing of childbearing

#### Fertility and the child as an important value

Women with a stronger conviction of a child as an important value wish to have it earlier. With each additional point on the subscale there is a 7% decrease in the chances of wanting a child later than in the next 5 years (aOR = 0.93, 95% CI: 0.90 – 0.96, Table 3a). This model explains 30.1% of the variance (Nagelkerke R^2^ = 0.301, p < 0.001).

**Table 3.**
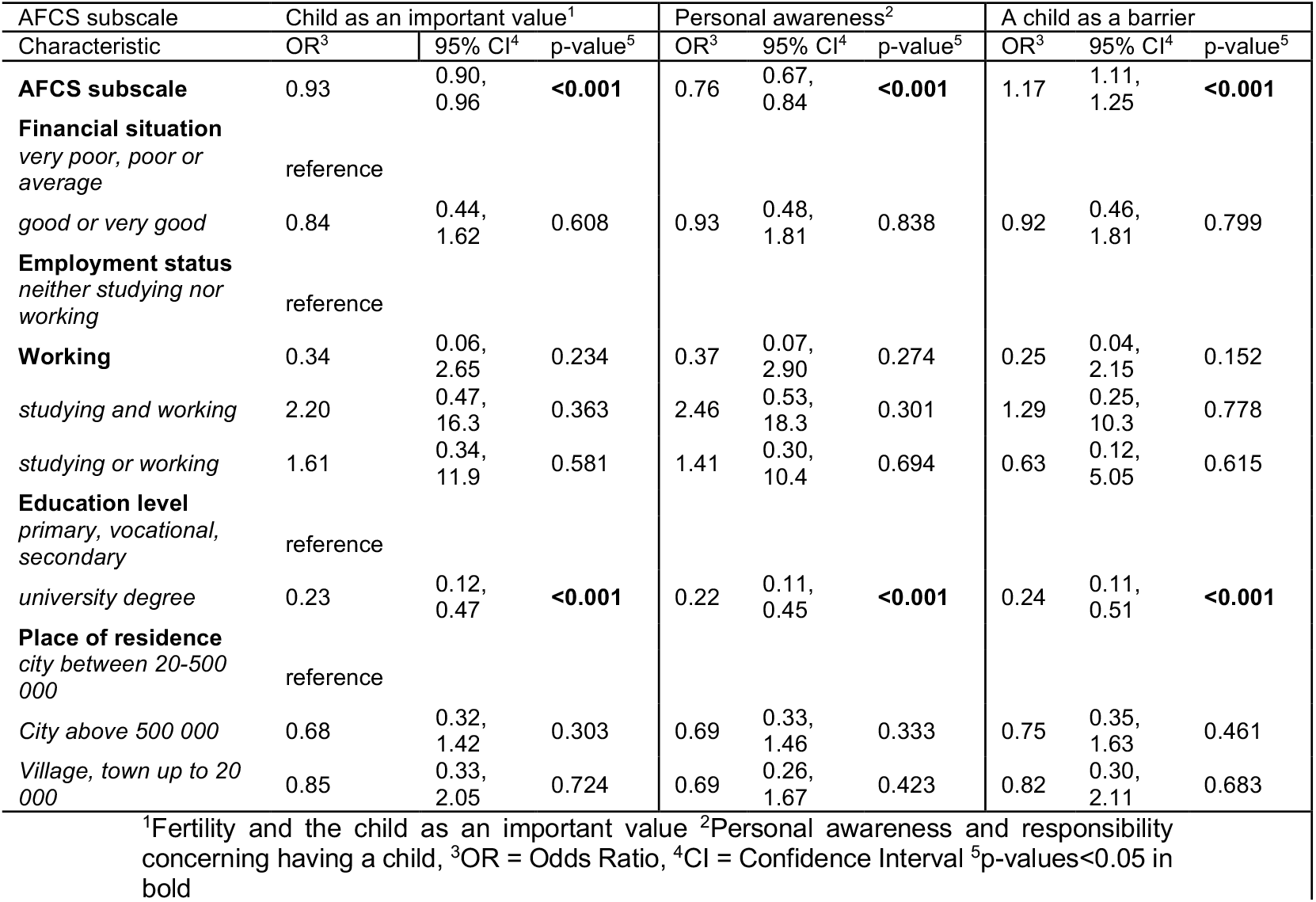
Timing of wanting to have a child and attitudes to the childbearing and fertility scale – results from multiple logistic regression models.

#### Personal awareness and responsibility concerning having a child

Women with higher personal awareness and responsibility concerning having a child wish to have it earlier. With each additional point on the subscale there was a 24% decrease of chances of wanting a child later than in the next 5 years (aOR = 0.76, 95% CI: 0.67 – 0.84, Table3b). This model explains 29.9% of the variance (Nagelkerke R^2^ = 0.299, p < 0.001).

#### A child as a barrier

Women who perceive a child as a barrier wish to have it later. With each additional point on the subscale there is a 18% increase in the chances of wanting a child later than in the next 5 years (aOR = 1.18, 95% CI: 1.12 – 1.25, Table 3c). This model explains 40.2% of the variance (Nagelkerke R^2^ = 0.402, p < 0.001).

## Discussion

Attitudes towards parenthood play an important role in shaping both the desire to have a child, and the timing of childbearing. Strong personal awareness and responsibility regarding fertility and the perceived value of children are positively associated with childbearing intentions– women with higher scores on those subscales have increased chances of wanting to have a child than women scoring lower on those subscales. However, as we hypothesise, a high score in a child as a barrier subscale increases the chances of not wanting to have a child and postponing reproductive plans. Women who perceive a child as a barrier are less likely to desire children. Our findings align with the results of Mynarska & Rytel, (2020), who analyse what childbearing motives are related to the fertility desires of Poles. Their research reveals that individuals who gave more importance to the negative aspects of parenting, such as time commitments, financial costs and energy demands were less inclined to have children. The comparison of these results highlights the stability of Polish women’s attitudes towards fertility despite the various crises to which Polish women have been exposed during the last five years (war and pandemic). Graham et al. (2015) found that perceiving parenthood as detrimental to freedom and career prospects significantly reduced the likelihood of motherhood. Perceiving a child as a source of emotional, economic and social costs also reduces fertility intentions (Kohlmann, 2002; Azmoude et al., 2017). Mills et al. (2011) highlight the role of changing values. According to the Second Demographic Transition Theory (Lesthaeghe, 1995; Van De Kaa, 1987), an increasingly individualistic family model has emerges. The family has become a less important value, and an increase in the number of divorces and cohabiting relationships has been observed (Van De Kaa, 1987). People of reproductive age increasingly prioritize self-fulfilment and personal development over parental norms and values (Mills et al., 2011). To achieve these goals, they seek higher education and satisfying careers, often viewing children as obstacles rather than enablers of these aspirations. These cultural preferences reflect what the Second Demographic Transition theory (Lesthaeghe, 1995; Van de Kaa, 1987) conceptualizes as a shift from traditional family formation toward individualistic, post-materialist values. Our findings give empirical support to this theoretical framework by demonstrating how perceptions of children as barriers are shaped not only by immediate life circumstances, but also by broader cultural narratives that promote personal autonomy, flexibility, and career ambition.

We show that women who perceive a child as a barrier want to postpone childbearing i.e., have a child later than in the next five years. Similar trends are observed by Dundar & Elverdi (2023) among Turkish women aged 20–30 and Lampic et al. (2006) among Swedish university students, who both indicate economic concerns and career planning as reasons for postponing parenthood. Similarly, Araban et al. (2020) emphasize the importance of socio-psychological factors, such as perceived social support, marital satisfaction and positive attitudes towards fertility in shaping fertility intentions for Iranian women. They are especially important for women intending to have children within two years (Araban et al., 2020). Lampic et al. (2006) and Verweij et al. (2020) also find that women wish to postpone motherhood until after the age of 30 driven by a desire to have a child at a later age, often based on a perceived ideal timing for first childbirth. These findings imply that while cultural contexts may influence the specific nuances of fertility attitudes, certain psychological and individual factors may have a universal impact, transcending cultural boundaries. Consistent with the Theory of Planned Behaviour (Ajzen, 1991) attitudes towards a given behaviour influence the formation of intentions. In our study, the perception of a child as a burden functions as a negative belief, thus lowering the intention to have children.

Women with greater awareness and responsibility regarding childbearing, and who highly value children, express a desire for earlier parenthood. Liefbroer (2005) argues that the decision to have a child is now a deliberate, conscious choice that significantly affects lifestyle: partnerships (an enhancement in quality and a sense of security within a relationship), and career/economic well-being (the potential for professional advancement). Another explanation may be related to the medical consequences of delaying childbearing (Schmidt et al., 2012). Women with higher levels of awareness and responsibility are likely to recognise the increased risk of medical complications such as infertility, a higher risk of foetal death, chromosomal abnormalities, multiple pregnancies and preterm birth associated with having children after 35 years of age (Schmidt et al., 2012; Nazaré et al., 2022). However, as Lampic et al., (2006) find, half of the university students surveyed plan to have children after the age of 35, and are unaware of the relationship between advanced maternal age and declining fertility. Thus, there is a high need for health education which could impact future reproductive decisions. The results can be interpreted through the lens of Expectancy and Value Theory (Eccles & Wigfield, 2002), which suggests that motivation to pursue a goal depends on both the value an individual places on the goal and their expectations of success. The findings of the present study indicate that women who place a high value on children and possess a sense of responsibility are more likely to perceive greater benefits and fewer barriers to having children. These motivational mechanisms help explain why women who perceive parenthood as meaningful and achievable express stronger and earlier fertility intentions.

Women who consider a child to be an important value express both a desire to have children and an intention of having them earlier. This finding aligns with McQuillan et al. (2015), who note that women who ascribe paramount importance to motherhood exhibit stronger parental intentions. Additionally, McQuillan et al. (2015) observe that socio-demographic factors, including relationship status, age and parity, influence fertility intentions indirectly through the salience of motherhood (readiness to become a mom). Positive attitudes towards children and motherhood increase the likelihood of becoming a mother (Graham et al., 2015). Moreover, the expectation that childbearing will bring significant personal benefits, such as maintaining a sense of youth and lower concerns about financial limitations, further enhances this probability (Liefbroer, 2005; Chappell, 2024; Tran et al., 2024).

Adding to the complexity of psychological incentives for childbearing, in recent years, a negative perception of parenthood has emerged. The term “intensive parenting” can be attributed to evolving standards in child-rearing practices, mark by mounting pressure to adhere to what is often termed (Nomaguchi & Milkie, 2020). This shift has led to an environment where parenting is perceived as increasingly demanding and costly. Moreover, research findings suggest that parents frequently reported diminished well-being in comparison to their childless counterparts, often experiencing a subsequent decline in well-being following the birth of a child (Hansen, 2012). Consequently, these factors have the potential to contribute to the formation of negative expectations about parenthood, which may, in turn, lead to a reduction in the desire to have children (Tran et al., 2024).

From a broader perspective, factors influencing reproductive desires can be grouped into three different levels: individual (microsystem), relational (mesosystem) and broader sociocultural influences (macrosystem) (Hashemzadeh et al., 2021). In addition, the specific temporal context in which a generation lives should be considered, as this is also a factor that influences fertility desires (chronosystem). This viewpoint is consistent with the findings of our study, as the timing of data collection – occurring during the pandemic and shortly after changes in abortion accessibility in Poland – may have influenced participants’ responses. However, recent studies suggest that crises like the pandemic and war may have a negligible or non-existent impact on fertility desires (Dundar & Elverdi, 2023; Golovina et al., preprint; Marcinkowska et al., 2025). Our findings suggest that personal considerations remain one of the strong determinants. Given the significant impact of fertility desires on total fertility rates, understanding childbearing perceptions among individuals of reproductive age is essential. This understanding is essential for unpacking the marked fertility decline observed in recent years. By employing a multi-theoretical approach this study offers a nuanced and comprehensive perspective on the motivations underlying childbearing decisions.

### Limitations and future directions

Data for the current study was collected during the COVID-19 pandemic, which may affect the results. However, research in this area is inconclusive. Some studies show that the COVID-19 pandemic affects reproductive desires, while others suggest that there is no such relationship (Dundar & Elverdi, 2023; Golovina et al., 2023; Golovina et al., preprint; Marcinkowska et al., 2025).

The study is cross-sectional – it is impossible to say whether women who experience changes in attitudes towards childbearing also change their fertility desires and actual reproduction. A future direction could be a longitudinal study that would include AFCS and then measure actual fertility or long-term changes in attitudes. Since our findings suggest that age may predict childbearing desires, it is likely that these desires are not stable but instead vary across different life stages— implying that the role of age in fertility intentions may shift over time.

Our study focuses solely on women. As fertility desires are frequently shaped by both of the possible future parents, further research on men could shed more light on the importance of their parenthood attitudes. Further research could also focus on combined couple studies and interaction between female and male fertility desires. Moreover, comparing the attitudes and desires of men and women cross-culturally could allow for pinpointing socio-demographic bases for childbearing decisions and gender differences.

## Conclusions

Overall, our study shows the importance of attitudes to fertility and childbearing when it comes to the fertility desires of women of reproductive age. As it is well known that socio-demographic determinants are impacting decisions about childbearing, it is important to further investigate what psychological motives are standing behind that or moderating that relationship. A clearer understanding of these motives will facilitate the development of targeted policy interventions to address the rapid decline in fertility rates.

## Data Availability

All data produced are available online at https://doi.org/10.17605/OSF.IO/7KJS5

## Acknowledgements

The authors would like to thank all participants of the study who voluntarily dedicated their time to participate in the current study.

## Data, scripts, code, and supplementary information availability

Data are available online: https://doi.org/10.17605/OSF.IO/7KJS5; Jurczak et al, 2026. Scripts and code are available online: https://doi.org/10.17605/OSF.IO/7KJS5; Jurczak et al, 2026.

